# Design, EHR Integration and Evaluation of Clinical Decision Support Workflows Driven by a Mortality Prediction Model to Promote Goal Concordant Care^*^

**DOI:** 10.1101/2023.02.14.23285926

**Authors:** Laura M. Roberts, Lorenzo A. Rossi, Finly Zachariah

**Affiliations:** City of Hope National Medical Center, Duarte, CA, USA

## Abstract

We present a clinical decision support (CDS) framework to promote goal-concordant care for cancer patients nearing end of life, enabled by a 90-day mortality prediction model. Design, workflow, and deployment for four model driven CDS tools are described alongside utilization patterns and detailed performance analysis of the first of such tools integrated into clinical practice: an alert promoting advance directive completion. The alert achieved a precision of 44.1% (95% CI: 39.6 - 48.8%) and a recall of 34% (95% CI: 30.3 - 37.4%) with 9.8% 90-day mortality prevalence over a period of 8 months. Our analysis shows that both precision and recall of the alert were significantly impacted by the underlying clinical workflows. Based on feedback, clinicians have accepted model-driven CDS as a legitimate means to prioritize patients for advance care planning interventions.

## Introduction

Patient-centered care has been championed as a cornerstone of high-quality healthcare for decades. Despite this, studies indicate that we still struggle to consistently engage patients in discussions about their values, perceptions of illness, and goals for care and often fail to identify the inflection points at which care plans must change to remain aligned with patients**’** expressed wishes.^1–3^ Resultant disparity between the care patients want and the care they receive has far-reaching ethical and economic consequences.

Timely, accurate prognostication is a key enabler of patient-centered care. While goals of care conversations empower patients to chart the course of their care journey with the help of their care team, changes in prognosis represent junctions at which the care plan must be revisited and potentially revised. Prognostication is challenging, influenced by complex biological, environmental, interpersonal, and temporal factors.^4, 5^ Various tools have been developed to support clinician prognostication, a growing number of which leverage artificial intelligence (AI) and electronic health record (EHR) data to automate mortality risk predictions.^6, 7^ To date, few AI mortality prediction models have been fully integrated with EHRs to work in real-time and suggest interventions within clinical workflows. Teams at NYU Langone and University of Pennsylvania Health System are among the first to do so and their work lends insight into the potential of these tools to support patient-centered care. At NYU Langone, use of a 60-day mortality model to suggest advance care planning (ACP) conversations and appropriate consults for hospitalized patients at risk for mortality significantly increased rates of ACP documentation.^8^ The University of Pennsylvania’s application of a 180-day mortality prediction model at eight medical oncology clinics resulted in a significant increase in ACP and serious illness conversation documentation not just in the high-risk population, but across all oncology patients.^9^

Motivated by similar aims to support broader efforts at our organization to improve goal-concordant care, we developed and prospectively evaluated a machine learning (ML) model to predict 90-day mortality via several non-interventional pilots. In parallel, we designed a technology-augmented operational framework (**Figure 1**) that leverages the model in an attempt to increase rates of high-quality advance directives (AD) and goals of care (GOC) conversations and appropriately direct limited Supportive Medicine (palliative care) resources. We deployed the model within the EHR at our organization then progressively introduced a series of ML-driven clinical decision support (CDS) workflows targeting subsets of the cancer population predicted by the ML model to be at moderate to high risk for 90-day mortality. Here we describe these CDS tools within their related workflows, explore findings around their use, and discuss lessons learned that may serve to inform other applications of AI-generated predictions in clinical workflows.

**Figure 1.**
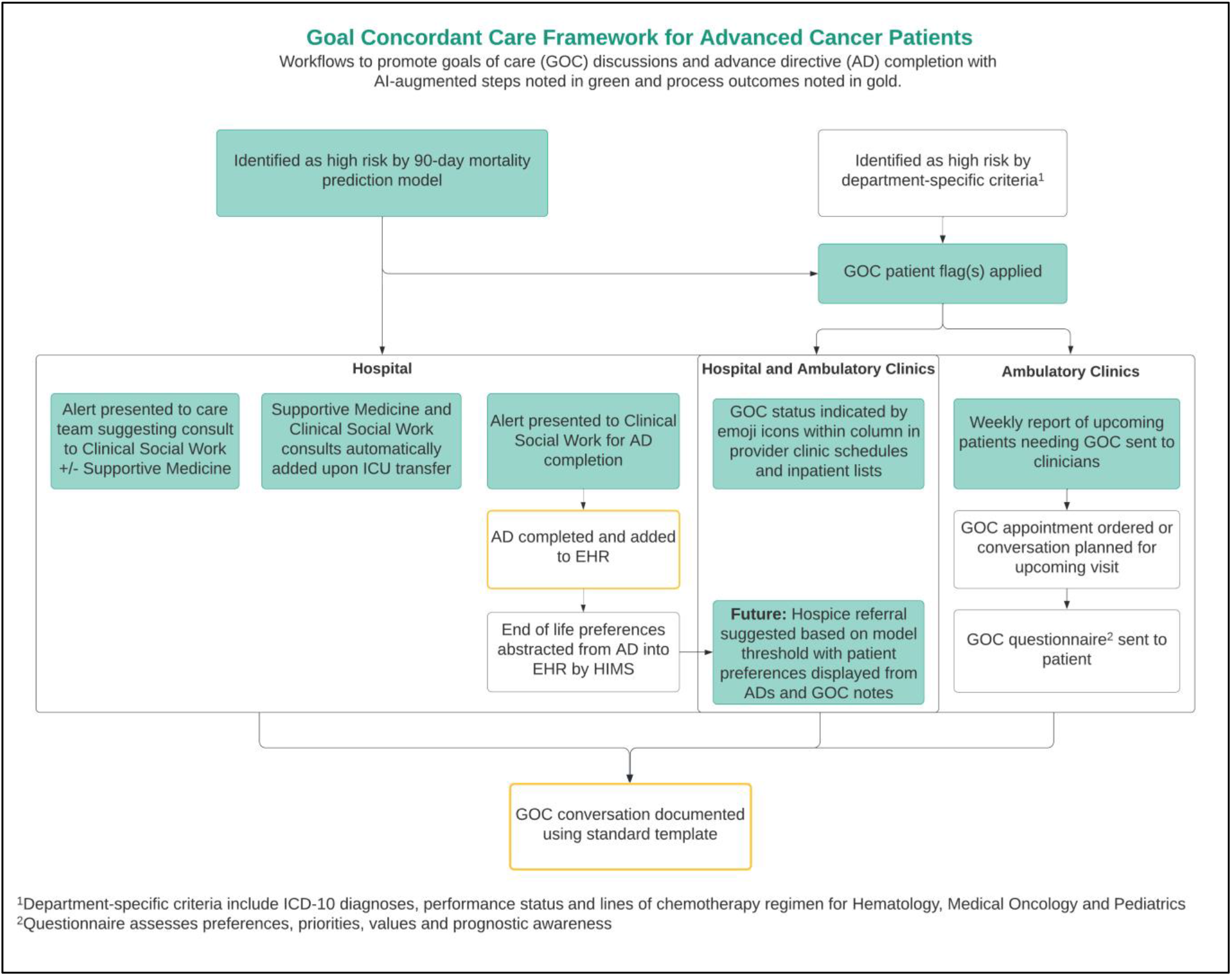
Goal-concordant care framework with AI-driven components denoted in green.

Given the significant upfront investment required to develop and comprehensively evaluate AI-driven mortality risk models, it is important to carefully and creatively consider how a single model might be applied to support clinicians and patients at different touchpoints across different settings. To the best of our knowledge, we are the first to apply a single model to multiple settings and clinician populations to promote an array of advance care planning (ACP) and goal concordant care interventions, including automation of Supportive Medicine (SM) and Clinical Social Work (CSW) consults for intensive care unit (ICU) patients.

## Methods

City of Hope National Medical Center is a comprehensive cancer center in Southern California that treats over 90,000 patients per year across one acute care hospital and 36 ambulatory clinics. As a member of the Alliance of Dedicated Cancer Centers participating in the Improving Goal Concordant Care (IGCC) initiative, City of Hope is highly invested in delivering compassionate, patient-centered care. We view technology as an enabler to help systematize and standardize the capture and honoring of our patients’ goals and values.

Our 90-day mortality prediction model combines features extracted from demographics, 6-month time series of lab test results and flowsheet data, and diagnoses. The features for the diagnoses are expressed as aggregations of Word2Vec embeddings of International Classification of Diseases level 9 (ICD-9) codes. We used the embeddings of ICD-9 codes trained by Choi *et al*. and shared in the related GitHub repository.^14^ The model is based on a gradient boosted tree binary classifier from the XGBoost library^15^ and was trained with EHR data from a cohort of 28,494 patients.^11^ Our implementation includes an explainability component^11^ to extract and weight the clinical factors that contributed the most to a prediction score from the related SHapley Additive exPlanations (SHAP)^10^. We evaluated different versions of the model over multiple silent prospective deployments based on data from City of Hope’s electronic data warehouse.^12^

We integrated the model into our organization’s Epic EHR via a custom real-time infrastructure in February 2021 then gradually introduced CDS tools that use the predictions to suggest different ACP interventions. The model predictions are triggered by results from a list of commonly ordered laboratory tests. Predictions are made across both hospital and ambulatory settings and repeatedly within the same encounter, unlike mortality models deployed in other organizations where predictions may only happen at the time of admission^8^. Inclusion criteria for specific patient subpopulations and alert suppression criteria are handled at the level of the CDS tools that reference the model. To date, prediction scores and explanations in the form of the top six clinical factors are displayed within two of these tools.

### Clinical decision support design

Goals of care conversations should happen early and often following an initial cancer diagnosis. We worked backwards towards this objective by prioritizing development of CDS targeting admitted patients predicted to be near end of life to maximize the impact of suggested interventions and build buy-in before expanding our framework to cover earlier timepoints and broader patient populations.

The CDS tools are summarized in **Table 1**. They include interruptive alerts as well as passive patient flags, icons within inpatient and clinic schedule columns and dynamic order presentation rules. Interruptive alerts were used judiciously and implemented only with strong support from clinical leaders and review by appropriate EHR governance, to introduce intentional pauses in clinical workflows at critical timepoints where patients nearing end of life were missing actionable documentation to guide goal-oriented care.

**Table 1.** Model-Driven clinical decision support (CDS) tools.

| CDS TOOL | TARGET POPULATION | GO LIVE | RECIPIENTS | PURPOSE |
| --- | --- | --- | --- | --- |
| **AD COMPLETION ALERT** | Admitted adult patients | February 2021 | Clinical social workers (CSWs) | Prioritize patients for advance directive completion and assess the need for GOC |
| **GOC ALERT RECOMMENDING CONSULTS** | Admitted adult hematology and medical oncology patients, excluding those within one month of transplant or CAR-T therapy | March 2021 | Inpatient care team providers | Consult CSWs for GOC conversation facilitation with optional Palliative Care/Supportive Medicine (SM) involvement |
| **SILENT GOC FLAG ALERT** | Ambulatory and admitted adult hematology and medical oncology patients | October 2021 | Inpatient/Outpatient care teams | Identify advanced cancer patients by model or departmental criteria who would benefit from GOC conversations |
| **ICU CONSULT AUTOMATION** | Admitted hematology and medical oncology adult patients being transferred to the ICU | January 2022 | Ordering providers | Engage SM and CSWs for GOC conversations |

### Advance directive completion alert

The first model-driven alert was launched in February of 2021 to increase AD completion and facilitate assessment for GOC discussions on moderate to high-risk patients. This AD completion alert (**Figure 2**.L) is presented to CSWs opening the chart of an admitted patient with a model risk score above a decision threshold. We set the threshold with the intent of achieving 50% precision (or positive predictive value), associated to medium mortality risk. While ACP and AD completion can benefit cancer patients at low risk of mortality, a higher threshold was chosen to balance opportunity with clinical capacity. Clinical leaders determined AD completion for patients meeting this threshold would be best facilitated by CSWs, making CSWs the logical recipients for the alert. CSWs are presented with multiple acknowledgement reasons to either indicate follow-up actions for AD completion or justify why an AD cannot be completed. The acknowledgement reason selected drives the interval for which the alert is prevented from appearing for the same individual on the same admission, ranging from three hours to 30 days.

**Figure 2.**
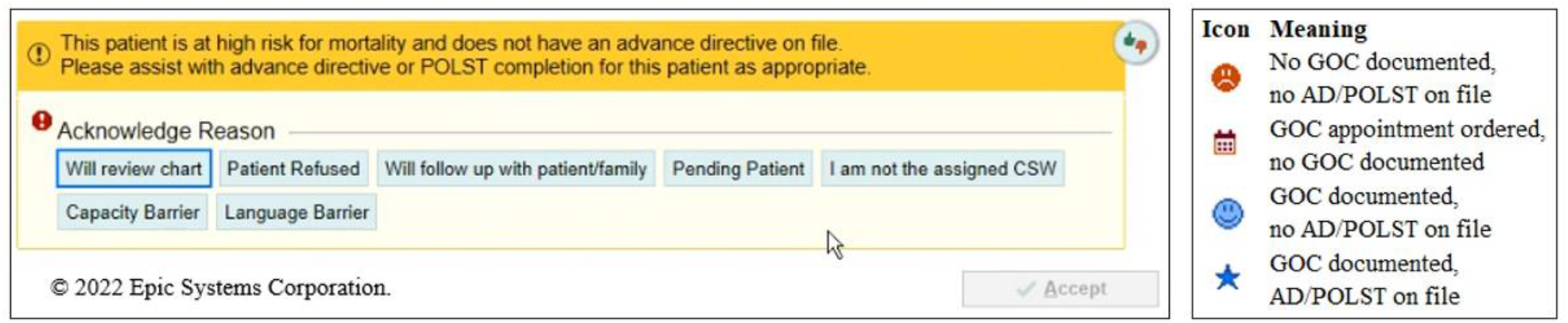
Advance directive completion alert (L) and icons presented in clinic schedules and inpatient lists (R)

### Goals of care alerts

Next, in collaboration with hematology and oncology, we implemented alerts to increase SM and CSW engagement in GOC conversations for seriously ill admitted patients. We set the decision threshold on the model predictions with the intent of achieving 60% precision. If the model “flags” an admitted patient without recent GOC documentation or SM consult, the alerts are presented to hematologists, medical oncologists, intensive care physicians, hospitalists, and advance practice providers (APPs) on the care team when they place orders. Based on pre-implementation feedback from hematologists, patients within one month of a bone marrow transplant or CAR-T infusion were excluded due to anticipated variability in lab results which providers were concerned would result in overestimation of mortality risk. Our evaluation of the model during the silent pilot confirmed a decrease in predictive performance for such categories of patients. Within the GOC alert, a 90-day mortality risk score and the top six clinical factors contributing to the score are presented and the provider is asked to indicate whether they believe 90-day mortality is very likely, likely, unclear, unlikely or very unlikely. The alert suggests a consult to CSW for a GOC conversation with an option to add a SM consult. If no orders are placed, the provider is required to indicate a reason using one of five pre-defined acknowledgement options. Selection of one of these options suppresses presentation of the alert for a set timeframe between five hours and 14 days. The alerts were set up to run silently without presentation to clinicians for the first two weeks to verify the volumes of notifications. For the first six months, prognostic information and order suggestions were presented to clinicians in two alerts presented simultaneously upon opening the chart. These were later combined into a single alert that displays upon signing orders for efficiency and better alignment with clinical workflows.

### Goals of care flag

In October of 2021, we extended use of the ML model to the ambulatory clinic setting by using it to trigger a silent (not presented to clinicians) alert that automatically applies a patient-level GOC flag for Hematology and Medical Oncology patients with advanced cancer predicted to be at high risk for mortality. Advanced cancer definitions were established by Hematology and Medical Oncology departments using ICD-10 diagnoses, lines of chemotherapy and performance status. Again, patients near bone marrow transplant and CAR-T infusion were excluded. In addition to the GOC flag, a new GOC column was added to the provider schedule and inpatient lists. This column automatically populates with one of four possible icons that serve as a quick visual indicator of GOC conversation and AD completion status for each advanced cancer patient with a GOC flag (**Figure 2**.R). While outside the scope of this analysis, supporting EHR build was also implemented to enable scheduling of GOC appointments, a patient-facing questionnaire and standardized documentation for GOC conversations.

### Supportive medicine consult automation for ICU transfers

Application of the ML model was further expanded in the hospital setting in January 2022 to dynamically present different consult defaults to providers transferring Hematology and Medical Oncology patients to the ICU. Prior to this time, SM and CSWs were consulted on most ICU transfers which was not always beneficial. To address this, the consult sections of ICU admission order sets were updated to present pre-selected SM and CSW consult orders if the patient in context is predicted to be at high risk for mortality. For patients not at high risk, SM and CSW consult orders are presented but not selected by default. Several weeks later, to cover cases where an ICU admission order set is not used, an update was made to present an interruptive alert to automatically add SM and CSW consults each time a standalone ICU transfer order is placed ordered for high-risk patients. Our evaluation of the silent pilots motivated the consult automation application because it showed that the model could achieve a higher recall on near death patients than the previous approach while maintaining the same volume of consults.

### Clinical implementation approach

We designed CDS and corresponding workflows in partnership with departmentally endorsed clinical champions, with input from clinical, operational, and technology-focused committees. We estimated volumes of alerts and predictive performance tradeoffs based on the silent deployments run throughout 2020. This information was shared with stakeholders to provide assurance that the proposed CDS tools would benefit patients, families, and the care team while respecting staff capacity and minimizing alert fatigue. We approached the initial development of each CDS tool by working with clinical champions to understand current state workflows and identify points at which CDS could be most seamlessly integrated to achieve intended outcomes. CDS solutions were then iteratively prototyped in Epic, presented to the appropriate stakeholder groups and adjusted per their feedback. Prototypes and formal requirements documentation were then handed off to the IT team for build. Clinicians were proactively engaged for user acceptance testing (UAT) and a final round of design and build adjustments were made to address their feedback. In parallel, CDS tools were socialized at department and division meetings and communicated to impacted staff. Post-implementation changes followed a similar path.

### Evaluation

We used Epic reporting tools to regularly monitor alert volumes and interactions following implementation. We met individually with clinicians experiencing the highest volumes of alerts to obtain their feedback and better understand unexpected alert interactions. Several group discussions were also conducted to provide opportunities for clinicians to raise issues and suggest enhancements. Information gleaned from regular monitoring and user interactions was used to further refine and enhance CDS tools post-implementation. In some cases, clinicians provided specific improvement suggestions which we reviewed with clinical champions, prototyped for consideration, and implemented if deemed appropriate. When feedback provided was more general or problem-based, we formulated solutions and presented these for consideration and decision. The AD completion alert was also formally evaluated using an anonymous survey sent to CSWs to assess usability and perceived impact the alert.

## Results

The counts of CDS presentations are summarized in **Table 2**. An alert may be triggered multiple times for a single patient if expected ACP interventions are not completed or documented in the chart within a given timeframe. The total number of alert instances is presented in the “Total Firings” column. We evaluated the volumes between the go-live date of each application and February 19^th^, 2022. Each CDS tool is driven by the model and targets different patient subpopulations and clinicians (**Table 1**), although with overlaps. The decision thresholds on the predictions vary across CDS tools as well. The silent GOC flag produced a relatively larger volume of alerts for the considered time frame due to higher volumes of ambulatory clinic encounters as compared to inpatient admissions. An in-depth study of the impact of the tools will be the focus of our Stage III manuscript.

**Table 2.** Presentation of the clinical decision support tools driven by the 90-day mortality model.

| CLINICAL<br>DECISION<br>SUPPORT TOOL | TIME FRAME | TOTAL<br>FIRINGS | DISTINCT<br>ALERTS | DISTINCT<br>PATIENTS | DISTINCT<br>PROVIDERS |
| --- | --- | --- | --- | --- | --- |
| AD<br>COMPLETION<br>ALERT | 2-16-2021 – 2-19-2022 | 2313 | 834 | 675 | 42 |
| GOG ALERT | 3-23-2021 – 2-19-2022 | 1031 | 517 | 412 | 112 |
| SILENT GOC<br>FLAG ALERT | 10-21-2021 – 2-19-2022 | NA | 1021 | 431 | NA |
| ICU CONSULT<br>AUTOMATION | 1-19-2022 – 2-19-2022 | 9 | 8 | 8 | 8 |

In the rest of this section, we report on the performance and usability of the AD completion alert because it has been live for the longest time span. Among the CDS tools based on the mortality model, the AD alert also has the lowest decision threshold and the highest volume of notifications per end user. Therefore, it is a good benchmark for volume and performance of the other CDS tools described in this manuscript.

### Performance of the advance directive completion alert

We considered the February 16^th^ – October 14^th^, 2021 period, choosing the stop date for the evaluation to account for delays in communication of death. The time frame chosen for the overall performance evaluation of the model in Stage I of the Evaluation Showcase was two weeks shorter.^12^ In the Stage I submission, we evaluated performance over all the predictions made by the model during the time frame^12^. In this work, we focused only on predictions for hospital patients without an AD (the target population for the AD completion alert) and considered only one prediction per patient encounter: either the first prediction above the threshold, or the last prediction before discharge.

**Figure 3**.L shows the weekly counts of AD alerts along with patients and providers involved. **Figure 3**.R shows the fraction for different subsets of acknowledgment reasons. We observed an increasing number of charts navigated by the CSWs since the go-live of the alert. As the plot shows, the same alert could fire multiple times until AD completion, depending on the chosen acknowledgment reasons. **Figure 4**.L shows the different counts of acknowledgment reasons. **Figure 4**.R shows the distribution of the volume of notifications across different CSWs. For two CSWs in supervising roles who navigate a larger volume of patient charts than non-supervisory CSWs, the AD alert fired over 90 times, i.e. on average 12 times a month.

**Figure 3.**
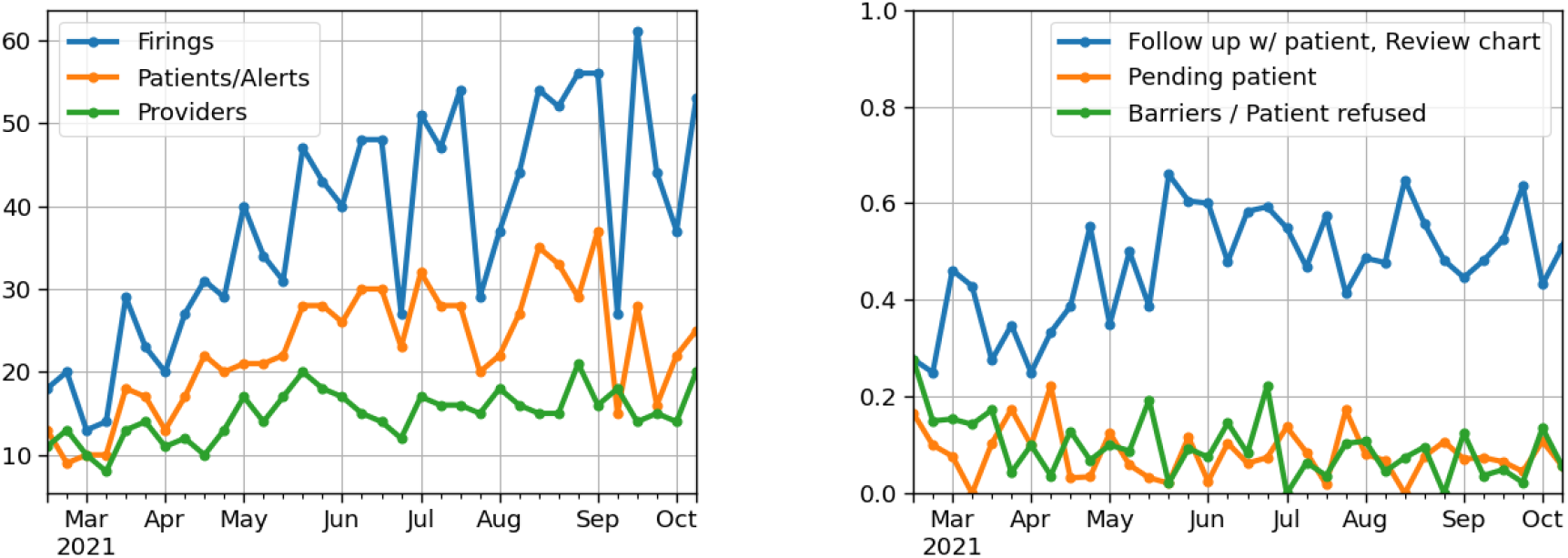
Weekly counts of firings, patients, unique alerts and providers for the AD completion alert (L). Weekly fractions of aggregated alert acknowledgment reasons for the same tool (R).

**Figure 4.**
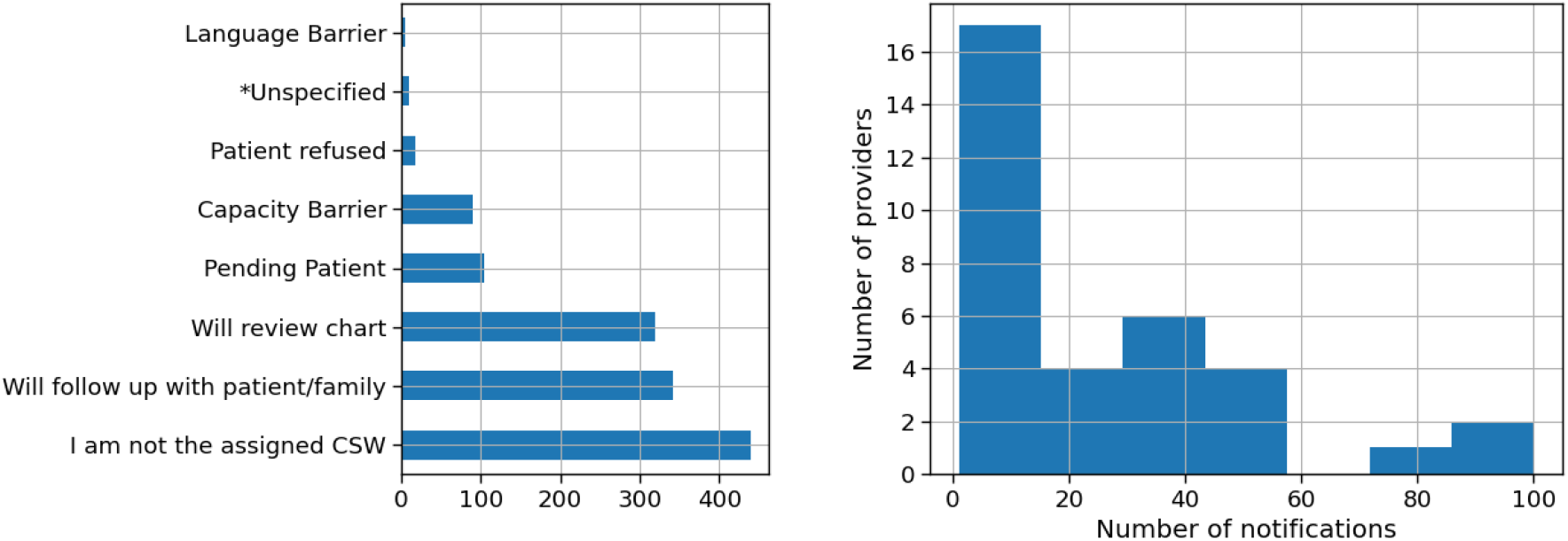
Counts of acknowledgment reasons chosen by CSWs for the AD completion alert (L). Distribution of the number of notifications over the number of providers over the Feb 15 – Oct 14, 2021 period.

The Receiver Operating Characteristic (ROC) and Precision-Recall curve (PRC) for the target subpopulations are displayed in **Figure 5**.L,R. The model achieved 0.83 (95% CI: 0.81 – 0.85) area under the ROC (AUROC), 0.4 (95% CI: 0.36 – 0.44) AUPRC and 0.08 Brier score with 9.8% 90-day mortality prevalence (n=6,324). AUROC measures the ability to discriminate between a positive (patients who die within 90 days of a prediction) and negative (patients who are still alive 90 days after a prediction) observation: a score of 0.5 indicates random guessing. AUPRC is a measure of performance with values between 0 and 1. The value of AUPRC associated to random guessing would be equal to the 90-day mortality rate in our population.

**Figure 5.**
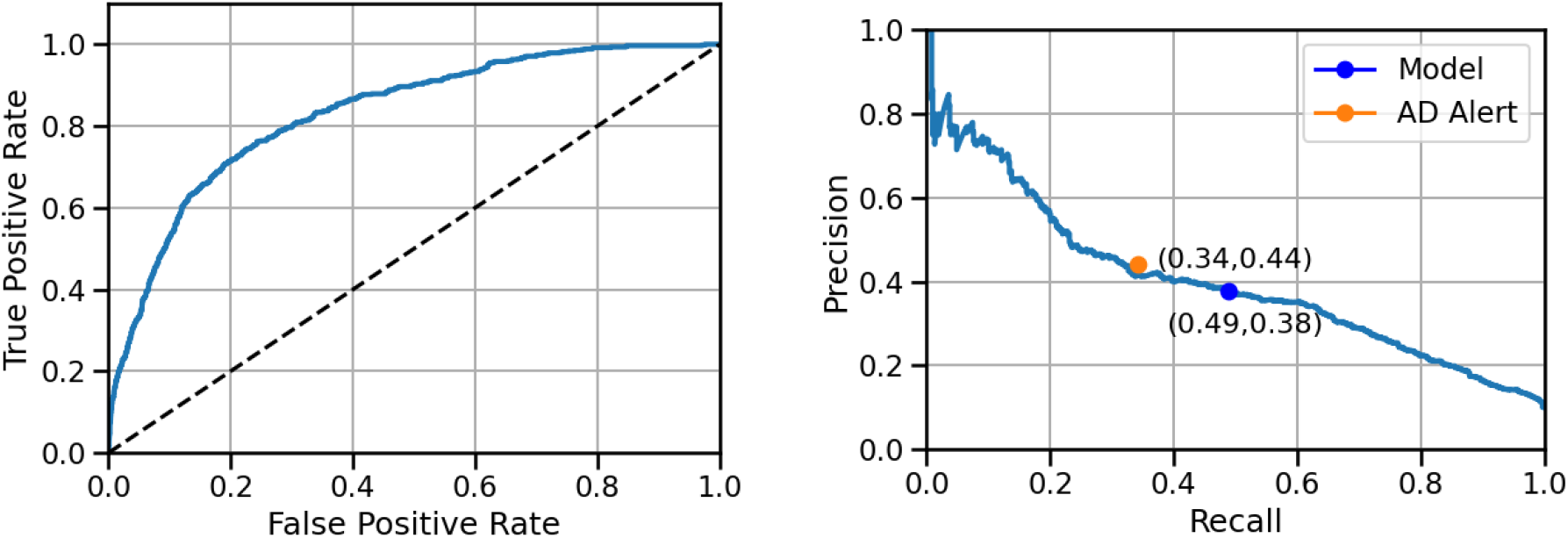
Receiver operating characteristic (L) and precision-recall (R) curves for the 90-day mortality prediction model over the target population for AD completion alert, with recall and precision for model and alert (R).

With the decision threshold we selected, the model achieved 48.5% recall and 38% precision. A prediction above the threshold is necessary but not sufficient condition to trigger the alert. The AD alert necessitates a CSW to navigate a patient’s chart as an additional condition for firing. CSWs do not have the capacity to navigate every admitted patient’s chart before discharge. This resulted in 34% recall and 44% precision for the AD alert, different from the model. Model and alert performance are summarized in **Table 3**, along with 95% confidence intervals (CIs). There is an overlap between CIs of precisions for model and alert. However, the evaluation of the difference between precisions of model and alert gave a 95% CI of (0.4-12.3%), indicating a statistically significant increase in precision (p < .05) for the alert. The difference in recalls was even higher: (95% CI: −17.5 - −11.9%).

**Table 3.**
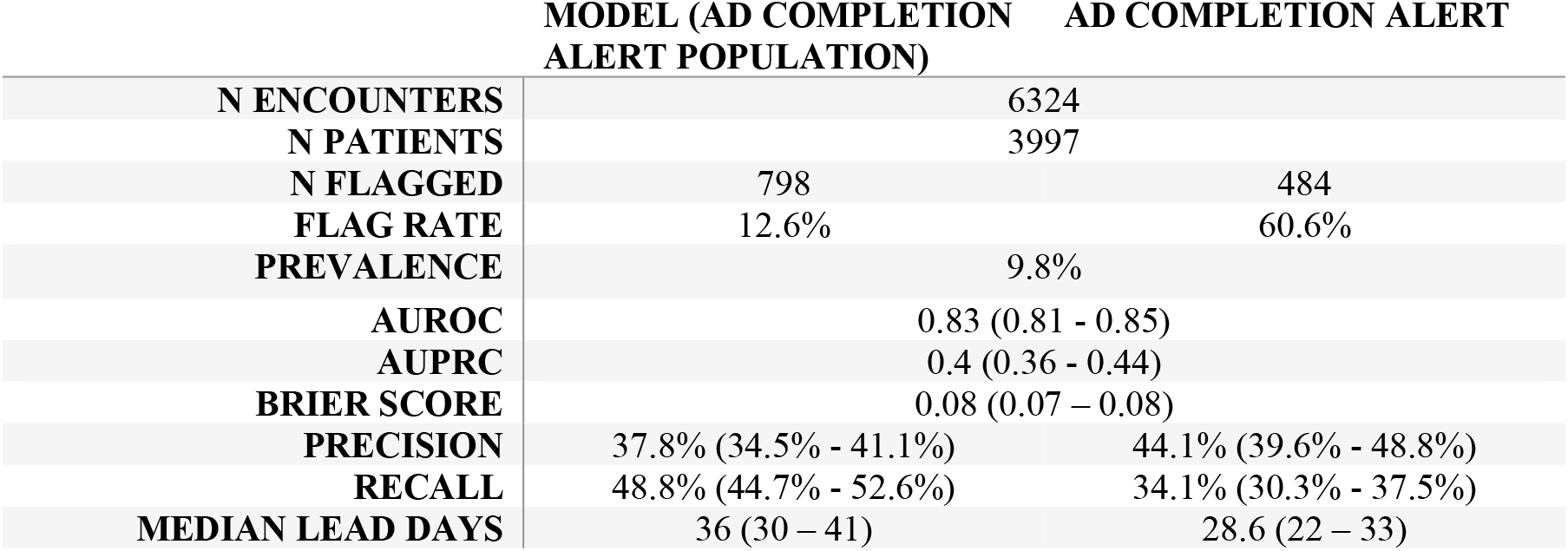
Performance of the model over the hospital patients without advance directive (AD) at admission between 2-16 and 10-14-2021. The column on the right shows the actual performance of the AD alert which comprises only a subset of the patients flagged by the model.

### Survey on advance directive completion alert

User feedback for the AD completion alert was obtained via an anonymized survey of CSWs who received the alert at least once. The survey consisted of six statements with mixed positive and negative framings and Likert scale response options, two workflow questions with pre-defined response options, and an opportunity to provide general feedback in narrative form. Of the 29 CSWs surveyed, 23 provided responses (79.3% response rate). 95.7% of respondents agreed or strongly agreed with the statement “It is simple to interact with and understand the alert.” None disagreed or strongly disagreed. When provided the statement “This alert helps me prioritize the patients I approach for advance directive completion”, 65.2% agreed or strongly agreed. Only 17.3% agreed or strongly agreed with the statement “I receive too many alerts for advance directive completion across all patients I see.” When asked “Would it be helpful to regularly receive a list of high-risk admitted patients to avoid potentially missing those whose charts you do not open?”, 43.5% responded “Yes, but I do not have bandwidth to consistently act on this information.” The remaining responses were split between “Yes” and “No, I always open the chart for my assigned patients so this would not provide me with any new information.”

## Discussion

The difference in volumes of distinct alerts and patients implies that there is a delay between receipt of a CDS notification and action taken on the suggested ACP interventions. It also suggests this delay tends to exceed alert suppression logic. For the GOC alert, we can attribute much of this to gaps in workflow and education. Initially, the GOC alerts were presented to clinicians upon opening a patient chart. Unless a clinician was already planning to sign other orders while in the chart, orders accepted from the alert were often queued up to be signed but then inadvertently discarded upon closing the chart. Upon recognizing this issue, the alerts were updated to be presented within the order signing workflow. Modest improvement in the volume of signed orders initiated from the GOC alert was observed following this change.

### Advance directive completion alert

CSWs prioritize seeing hospitalized patients at highest risk, including patients identified from clinical huddles and referrals. We observed that the CSWs navigated the charts of about 61% of the instances identified by the model **(Table 3)**. The most obvious consequence of this is a reduction in recall for the alert to 34.1% from the 48.8% achieved by the model. The reduction in median lead days between model and alert is due to the gap between the time of flagging and the navigation time. We expected a precision of around 50% for the threshold we chose based on performance of the pilot version of the model in 2020 but actual precision of the model on the target population was 38%. The operating precision associated with the alerts was higher: 44%. During the 2020 prospective pilot we observed a prevalence of 15% which is higher than 9.8% prevalence observed thus far. Note that AUROCs for model post go-live and 2020 silent pilot were remarkably close with only a 1% difference. We believe that higher 2020 prevalence was mostly associated to inpatient population shifts due to the COVID-19 pandemic and that the subsequent drop in prevalence was one the factors that contributed to overestimating the precision of the model. The lift in precision of the alert seems associated with the fact that CSWs are required to prioritize the most seriously ill patients. While the alerts had higher precision than the model, the recall, not surprisingly, was lower due to fact not all the charts of the flagged patients were navigated.

Several months after implementation, CSWs reported a challenge with the alert design. For newly assigned patients, the alert is presented before their chart can be reviewed to assess. As a workaround, CSWs were selecting “I am not the assigned CSW”, resulting in the high utilization of this option as pictured in **Figure 4**.L. This response prevents the alert from reappearing for the receiving CSW for the next seven days. To address this, a new “Will review chart” option was added that allows CSW to temporarily proceed past the alert, suppressing reappearance for just three hours.

The AD completion alert survey results indicate that overall, CSWs feel the alert is easy to use and serves the intended purpose of assisting in prioritization of patients for AD completion without exceeding their capacity. Feedback from almost half of respondents indicating that eliminating the chart navigation requirement to instead provide notification for *all* high-risk patients would exceed their capacity reinforces our capacity-conscious design approach. A similar survey to assess clinician perspectives for the GOCs alerts is currently underway. In addition to the survey results, adoption of the model as the primary pathway for identification of patients for ACP interventions suggests broad user acceptance. CSW, Hematology and Medical Oncology departments recognize the prognostic model as a valid way to identify patients for ACP interventions and have ingrained it in their care delivery processes.

### Key lessons learned

Operationalizing use of this ML model at our organization was an institutional learning experience. Continued collaboration between the project team, IT department and clinicians surfaced improvement opportunities and enabled swift adaptations in response. Here we describe several experiences of this process in action and the insights gained as a result.

Shortly after implementation, we discovered a discrepancy between the volume of SM consults accepted from GOC alerts and the total volume of signed consult orders. Further investigation led to a realization that orders accepted from alerts often go unsigned. Learning this alongside our clinical partners allowed us to avoid a scenario where the alert was deactivated due to perceived failure to achieve expected process outcomes. Instead, there was immediate operational support to quickly pivot from presenting the alerts upon opening the chart to presenting them within the ordering workflow. This experience heightened our awareness of the importance of understanding EHR utilization patterns prior to deploying new CDS tools and has prompted broader evaluation of provider interactions with all alerts containing order suggestions.

Operational readiness was both an enabler and barrier to successful CDS implementation. The project team spent substantial time meeting with departmental leaders to explain the model, share performance findings and CDS prototypes and review detailed projections around anticipated volumes of interruptive alerts and SM ICU consults. While time-consuming, we believe this work was instrumental in building support for eventual implementation of these tools by providing a forum for collaborative, data-driven cost/benefit discussions. Similarly, engaging CDS users in UAT and post-implementation feedback meetings and directly incorporating their suggestions as enhancements surfaced use cases that likely would not have been identified by the project team, while also reinforcing appropriate use of CDS tools and conveying a commitment for continued improvement. Conversely, inpatient providers frequently expressed discomfort around initiating GOC conversations in the acute care setting, due to patient acuity and widely held beliefs that primary oncologists and hematologists who typically have a longstanding relationship with the patient are better positioned to facilitate these discussions. While this feedback helped to accelerate implementation of the GOC flag and supporting ambulatory workflow, engagement of clinical leaders to set clear expectations around shared ownership of GOC conversations in advance of implementation may have helped to reinforce the role of inpatient providers in initiating these important conversations in the acute care setting.

When operating in dynamic healthcare environments, maintaining a holistic view of the entire ecosystem of ACP and end-of-life workflows and evolving these to maintain a streamlined clinician experience is crucial. Initially, pre-existing CDS to suggest SM engagement for patients manually identified for GOC discussions using departmental criteria was not deactivated in parallel with the introduction of the GOC alerts and GOC patient flag. This resulted in confusion and duplication of work by the model and APPs. This has since been addressed and has enabled a shift in clinician focus from the administrative task of identifying patients for GOC conversations to improved capture of information important for overall patient care, such as ECOG performance status. Mindfulness of the end-to-end clinician experience with constant re-evaluation of CDS is required to craft a unified user experience.

### Limitations

Recent implementation, frequent design changes, informal feedback collection methods and delays in recording of external deaths limit the volume of data available for analysis. This is further compounded by the fact that CDS was deployed broadly across entire clinician and patient populations rather than designing randomized controlled trials (RCTs), limiting the ability to draw direct cause-and-effect conclusions around patient outcomes. Recent work demonstrates the feasibility of using rapid RCTs to evaluate CDS embedded in EHRs and opportunity exists to evaluate future CDS using these methods.^13^

### Future work

The 90-day mortality risk prediction model is now an integral component of efforts to provide goal-concordant care at our organization. We are building on this work by developing additional CDS interventions, including alerts to suggest hospice education or admission based on specific risk thresholds and documented patient preferences. We are also developing workflows that will leverage the model to support recruitment for studies involving post-mortem tissue collection. The need to improve patients’ end of life experiences continues to be the driving force behind this work. As such, our future roadmap also includes constant evaluation of the impact of model-driven CDS in relation to patient outcomes. This will be the focus of our Stage III submission.

## Conclusion

By working closely with clinical champions, we introduced and refined a diverse set of clinical decision support (CDS) tools for advance care planning in our organization’s EHR, driven by a custom prognostic machine learning (ML) model. The CDS tools are used by clinicians in both hospital and ambulatory settings to identify patients to prioritize for goal-oriented care. In-depth analysis of the performance of one of these tools, an alert to promote advanced directive completion, revealed a loss in recall (95% CI difference: −17.5 - −11.9%) but an increase in precision (95% CI difference: 0.4 - 12.3%) following application in clinical practice, if compared with the same metrics for the ML model applied to the alert target population. The loss in recall was due to known capacity limitations of the clinical social workers. On the other hand, we did not anticipate a significant increase in precision, resulting from CSWs prioritizing seriously ill patients and patients they were consulted for. Feedback from the clinical social workers receiving the alert indicates it is understandable and perceived to be a useful and manageable addition to their clinical practice. Ongoing partnerships between institutional stakeholders, and the project team continue to be critical for adoption and optimization of AI models and downstream CDS tools to support institutional efforts to deliver high quality, value-aligned care to patients at the end of life.

## Data Availability

The data associated with the current study is not publicly available as the data are not legally certified as being deidentified. Summary data may be available by contacting the corresponding author.

